# Impact of climatic parameters on COVID-19 pandemic in India: analysis and prediction

**DOI:** 10.1101/2020.07.25.20161919

**Authors:** Namrata Deyal, Vipin Tiwari, Nandan S. Bisht

## Abstract

The COVID-19 is spreading very fast globally and various factors of it have to be analysed. The aim of this study is to analyze the effect of climatic parameters (Average Temperature (AT), Atmospheric Pressure (AP), Relative Humidity (RH), Solar Radiation (SR) and Wind Speed (WS)) on the COVID-19 epidemic during 25 March 2020 to 15 June 2020 in most affected states of India i.e. Maharashtra, Delhi and Tamilnadu. We quantitatively establish the correlation between these parameters by using Kendall & Spearman rank correlation test. The results indicate that the numbers of cases are highly correlated with the AT (r^2^ > 0.6, *p* < 0.001) in Delhi where as a moderate correlation (r^2^ > 0.6, *p* < 0.001) has been estimated for Maharashtra and Tamilnadu. Similarly, an intermediate range of correlation coefficient has been observed for other climatic parameters. A comparative study of climatic parameters in the current COVID-19 period with previous two years (2018-2019) has been carried out. Corresponding results imply a substantial trend for all three states. The range of climatic parameters have been found corresponding to maximum number of cases results as AT (25∼ 40 ° C), RH (40∼70%), AT (740∼965 mmHg), SR (200-250 W/mt ^2^) and WS (.5∼14 m/sec). Time series analysis depicts that the number of cases and mortality are increasing rapidly. COVID-19 epidemic peak has been predicted by SIR model for capital of India (New Delhi) and it would be around October 2020. The outcomes of this study will be helpful for the containment of COVID- 19 not only in India but globally.

## Introduction

COVID-19 has been declared as a worldwide pandemic by World Health organization (WHO) on March 11, 2020 (Cucinotta et al., 2020). Globally, the first COVID-19 case was reported on December 31, 2019 in Wuhan (China) (Zhu et al., 2020, Guan et al., 2020 Li et al., 2020, Deepak et al., 2020). At present, it has affected around 80% of world population and still growing at decent rate (https://www.worldometers.info/coronavirus). Investigation on COVID-19 recognized that its transition occurred by respiratory droplets, as well as human to human transition (Ge et al., 2013, Huang et al., 2020, Vandini et al., 2013). The common symptoms of COVID-19 infected patients are fever, cough and respiratory disorders (Holshue et al., 2020). In worst conditions, it might results as serious health issues like kidney failure, pneumonia which might cause death of patients (Wang et al., 2020, Ten et al., 2005, Perman,2020). The major concerns about COVID-19 are its tremendously growing cases and vulnerable community transmission in world. In addition, no vaccination of COVID-19 has been officially reported till date. Therefore, adequate precautions and preliminary research work on the factors affecting the spreading of COVID-19 might be helpful for development of vaccination process of COVID-19. Recent studies suggest that the spreading of COVID-19 is highly correlated with the atmospheric factors such as temperature, humidity etc (Ma et al., 2020, Chen et al.,2020, Qi et al.,2020, Wang et al.,2020). It has been reported that abrupt change in climatic conditions and population might be responsible for virus transmission (Rockloy et al., 2020, Sohrabi et al., 2020, Dalziel et al.,2018, Jaiswal et al.,2015, Hansel eta al., 2016). Conflictingly, few studies are not accounting meteorological parameters as carriers of transition of COVID-19 (Jamil et al.,2020, Mollalo et al.,2020, Shi et al., 2020). Another study indicates that the temperature, humidity can be responsible of transmission and existence of SARS-COV virus [(Bashir et al., 2020, Shi et al., 2020 b, Tan et al., 2020, Yuan et al., 2020). However, limited studies have been carried out in context of COVID-19 and climatic factors. *Tosepu et al*. have studied the correlation between weather and COVID-19 in Jakarta (Indonesia) in earliest stage of COVID-19 (Tosepu et al,. 2020) and predicted a connection between climatic factors (rainfall, temperature, humidity) and COVID-19 transmission cases. A few numbers of such works have also been reported but all these studies have been performed at the earliest of COVID-19 transmission and incorporate only limited data set (up to April 2020) (Ahmadi et al.,2020, Gupta et al,. 2020, Poole et al.,2020). Further, it is expected that this correlation also depends on geographical conditions of study area. So far, such reported studies are only limited to European countries (Briz et al., 2020, Sajadi et al., 2020). To best of author’s knowledge, no such study has been carried out for south Asian countries till date.

In this paper, we study the effects of varying climatic parameters (CPs) on the spread of COVID- 19 from 15 March 2020 to 15 June 2020 in India. The aim of this study is to analyse journey of COVID-19 in India and forecast the effect of COVID-19 on climatic conditions in subsequent times. Moreover, to obtain detailed analysis of the COVID-19, we have emphasized our study to three most COVD-19 affected states of India i.e. Maharashtra, Tamilnadu and Delhi. In addition, the strategies such as nationwide lockdown implemented by Indian government to reduce COVID-19 spreading have been quantitatively evaluated in climatic framework. In India, the first confirmed COVID-19 case was reported in Kerala on January 30, 2020 (https://www.mohfw.gov.in). The total confirmed cases has been raised up to 3,54,065 within five months in India, which is the highest number of confirmed COVID-19 cases in Asia and fourth highest in world as on 15 June 2020 (https://www.worldometers.info/coronavirus). It implies that COVID-19 is drastically spreading among 1.3 billions of people in India. Out of the total confirmed cases, 1, 86,935 patients have been recovered and total 11,900 deaths in country till mid June 2020 (https://www.covid19india.org). As anticipation, nationwide lockdown was imposed by Indian government in five phases. During this lockdown period, all social activities like transport, industries, shopping malls etc. had been strictly prohibited in India.

## 2. Methodology

### 2.1 Study area

India is the second-highest populated country (13‪10^8^, 17.7% of worldwide) after China located at north of the equator between 8°4’ north to 37°6’ north latitude and 68°7’ east to 97°25’ east longitude. It stands as the seventh-largest country in the world, with a total area of 3.28 x10^6^ km^2^. It is surrounded by Arabian Sea (in west), Indian Ocean (in south), and Bay of Bengal (in east). The north-east region of India has been covered with the Himalayas. Fig.1 shows the study area. The study further focuses on the three most crucial states of India in COVID-19 transmission i.e. Delhi, Maharashtra and Tamilnadu. These are the three major states of India.The population of these states are 1.8×10^7^, 1.23×10^8^ and 7.7×10^7^ respectively as on 2019 (Census 2011; http://census2011.co.in). Most importantly, these three are most affected states of India from COVID-19 as on mid June 2020.

### 2.2 Data collection

The digital dataset for the COVID-19 in India has been obtained from Ministry of Health and Family Welfare (MoHFW) [https://www.mohfw.gov.in/] and official website of covid-19 [https://www.covid19india.org/]. The data related to CPs (AT, RH, AP, SR and WS) are authentically retrieved from official online portal of Central Pollution Control Board (CPCB) [https://www.cpcb.nic.in/]. Retrieved data of CPs and COVID-19 from the sources were not distributed normally. Therefore for estimating correlation coefficient between parameters we use Kendall and spearman rank test. Further SIR model has been used for prediction of epidemic of COVID-19 (Weisstein, Eric W). For interpretation of results and graphics we utilized machine learning technique.

## 3 Results and discussion

The spatial distribution of COVID -19 parameters (Positive cases (PC), recovery and Death(DT)) during the lockdown in India is shown in fig.2. The whole lockdown period has been categorized in two phases i.e. period-I (from 25 March 2020 to 3 May 2020) and period- II (from 4 May to 15 June). Fig. 2 clearly indicates that Delhi, Maharashtra and Tamil Nadu are the most affected states of India from COVID-19 pandemic till mid June 2020. The additional details of COVID-19 parameters during the different lockdown phases are mentioned in (Table.1). On observing table.1 carefully, it is clear that PC and mortality increases rapidly all over India during lockdown period. One more important observation is that the cases are still increases as on June 2020 and the saturation state of COVID-19 cases still not achieved in India.

**Table 1:**
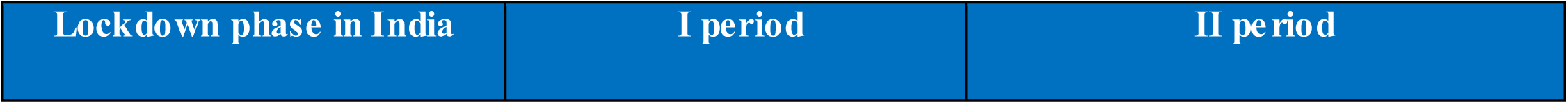
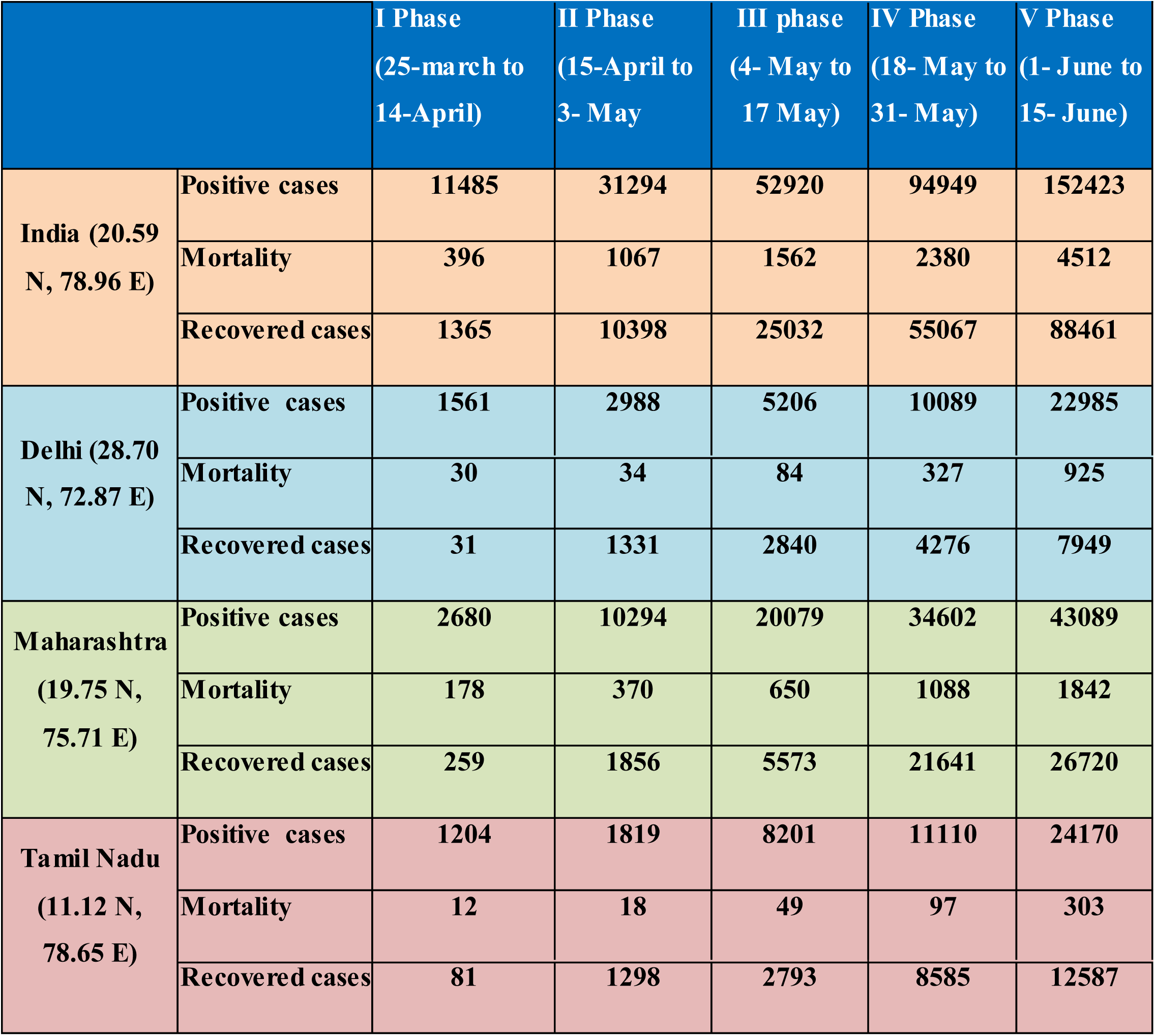
Details of COVID-19 parameters during different phases of Lockdown.

The variation in CPs (24hrs average value) of during the period of three months (25^th^ April to 15^th^ June) has been observed for previous three years (2018-2020) for three states of India i.e. Maharashtra, Delhi and Tamilnadu illustrated in fig.3 (a),(b),(c). We have observed similar trends in 2020 as compared to 2018 and 2019. However, a unique quantitative variation in CPs has been noticed for year 2020. AT significantly reduces with respect to previous years in the range of (5.2% ∼ 10.4%). Moreover, the RH represents ascending trend in all three years with relative difference (2.4% to 40%). It is an intuition that this variation is correlated with the COVID-19 pandemic up to some extent. Moreover, the study of variation of CPs has been carried out during lockdown period in Maharashtra, Delhi and Tamilnadu. Corresponding parameters have been tabulated in (Table.2). It depicts that the implementation of lockdown is a factor for considerable variations in CPs and air quality.

**Table 2:** Relative difference between climatic parameters for year 2018-2020(25 March to 15 June)

|  | Delhi |  |  | Maharashtra |  |  | Tamil Nadu |  |  |
| --- | --- | --- | --- | --- | --- | --- | --- | --- | --- |
| Climatic Parameter | Avg.2020 | Relative difference (%) |  | Avg.2020 | Relative difference(%) |  | Avg.2020 | Relative difference(%) |  |
|  |  | 2018-2019 | 2019-2020 |  | 2018-2019 | 2019-2020 |  | 2018-2019 | 2019-2020 |
| Average Temperature | 29 | 9.5 | 10.45 | 31.64 | 5.2 | 5.99 | 28.9 | 8.6 | 7.4 |
| Relative Humidity | 50.59 | 2.4 | 38 | 68.07 | 2.4 | 2.4 | 66.27 | 11.54 | 21.09 |
| Atmospheric pressure | 982.38 | 18 | 0.17 | 734.68 | 40.18 | 40.19 | 749.8 | 33.85 | 31.8 |
| Wind Speed | 0.78 | 92.3 | 41.02 | 0.113 | 21.9 | 22.2 | 9.482 | 77.28 | 75.84 |
| Solar Radiation | 189.23 | 13.8 | 17.6 | 158.28 | 29.9 | 60.1 | 242.72 | 7.7 | 10.2 |

CPs (AT, AP, RH, SR, and WS) play an important role in stability of environment. To quantify the correlation between COVID-19 spread and CPs, we have statistically analyzed the data and estimated correlation coefficient between CPs and COVID-19 parameters by using Spearman rank test and Kendall test as shown in (Table.3). It was observed that number of (PC) was highly correlated with AT for Delhi (r^2^ > 0.6) whereas moderate correlation is observed for Tamil Nadu and Maharashtra (r^2^ < 0.6) with significant level of 0.1 % (p < .001

**Table 3:** Correlation coefficients between climatic parameters with COVID-19 parameters.

| Test | Climatic Parameters | Delhi |  | Maharashtra |  | Tamil Nadu |  |
| --- | --- | --- | --- | --- | --- | --- | --- |
|  |  | Positive cases | Mortality | Positive cases | Mortality | Positive cases | Mortality |
| Spearman<br>Correlation<br>Coefficient | Average Temperature | 0.821 | 0.753 | 0.099 | 0.071 | -0.550 | -0.536 |
|  | Relative Humidity | 0.054 | -0.209 | -0.438 | -0.473 | 0.615 | 0.560 |
|  | Atmospheric Pressure | -0.842 | -0.736 | -0.784 | -0.797 | -0.799 | -0.774 |
|  | Wind speed | 0.326 | 0.047 | 0.421 | 0.413 | 0.733 | 0.722 |
|  | Solar Radiation | 0.334 | 0.270 | -0.297 | -0.294 | -0.329 | -0.413 |
| Kendall<br>Correlation<br>Coefficient | Average Temperature | 0.635 | 0.635 | 0.081 | 0.056 | -0.390 | -0.398 |
|  | Relative Humidity | 0.030 | -0.172 | -0.310 | -0.334 | 0.465 | 0.423 |
|  | Atmospheric Pressure | -0.646 | -0.563 | -0.580 | -0.602 | -0.559 | -0.559 |
|  | Wind speed | 0.200 | 0.040 | 0.263 | 0.256 | 0.529 | 0.550 |
|  | Solar Radiation | 0.214 | 0.187 | -0.176 | -0.180 | -0.228 | -0.302 |

In addition, the significant correlation between RH and PC was observed for the three states i.e. positive correlation (r^2^ > 0.6, *p* < 0.001) and intermediate correlation for Delhi (p > 0.1) and Maharashtra (r^2^ < 0.6). However, SR is not significantly correlated with COVID-parameters. Meanwhile, it is observed that WS affect COVID-19 cases as the corresponding correlation coefficient is positive. In brief, the obtained statistical results for most of the CPs were found very significant with (*p* < 0.001) in India. It is further observed that the mortality rate is growing with time. The CPs were shows a moderate correlation with DT. To have more clear insight, the scattered correlation matrices of CPs and COVID-19 parameters for Delhi, Maharashtra and Tamil Nadu shown in fig. 4(a), 4(b) and 4(c). It is observed that the maximum PC have been reported within the AT range (25∼40 ° C). Similarly, the most frequent ranges for other CPs corresponding to most number of PC are 40∼70%, 740∼965 mmHg and 200∼250 W/mt ^2^ for RH, AP, SR respectively. Particularly, a wide range for WS (.5∼ 14 m/sec) has been calculated for having maximum no. of PC from Delhi to Tamilnadu and varying with latitude. In lower latitude the no. of PC are high within the range of WS (10∼14 m/sec) although for northern region at higher latitude, it is observed (.5∼1.5 m/sec). This lead to conclude that the areas experience such climatically condition have been mostly affected by COVID-19.

Figure 5 represent the time series of no. of PC and mortality during lockdown period (25^th^ March - 15^th^ June) for three crucial states of India i.e. Delhi, Maharashtra and Tamil Nadu. The corresponding plot indicates that variation in PC and mortality cases increase exponentially with respect to time. Fig.6 provides a prediction of the number of COVID-19 cases in subsequent times in Delhi. The epidemic peak for COVID-19 in Delhi has been predicted on October 2020 (Weisstein, Eric W).

## Conclusion

The CPs (AT, RH, AP, SR and WS) are one of the crucial factors for in COVID-19 dissemination in India. On analyzing the available COVID-19 dataset, we have observed that the number of COVID-19 cases are still growing significantly despite of imposing containment strategies i.e. lockdown in India. It clearly indicates that there are some other factors (CPs) other than social activities which are influencing COVID-19 growth in India. It has been also observed that growth rate of COVID-19 is highest in three states of India i.e. Maharashtra, Delhi and Tamilnadu. In Delhi, the number of cases are positively correlated with AT (r^2^ > 0.6) whereas the correlation is moderate for other two states. Similarly, all other CPs (AP, RH SR and WS) show critical correlation with transmission of COVID-19 in India with 0.1% of significance level (*p* < 0.001). Further, the substantial variations of CPs have also been observed in last three years. On comparing the AT (daily average of 24 hours) of previous three years (2018-2020), a unique trend (decreasing comparatively) is observed. Moreover the range of CPs have been found corresponding to maximum number of cases results as AT (25∼40 ° C), RH (40∼70%), AT (740∼965 mmHg), SR (200-250 W/mt ^2^) and WS (.5∼14 m/sec). This signified that for the maximum transition of COVID-19, a susceptible weathers condition is required. In addition, the epidemic peak (highest number of cases) in New Delhi (capital of India) has been predicted around October-November 2020. It implies that COVID-19 transmission decreases in winter in India which compliment the results obtained from correlation test.

Despite of the enthusiastic outcomes of this study, there are further specific factors (people emigration, various government policies for containment etc.) which must be considered to obtain a more accurate prediction of COVID-19 epidemic parameters. Nevertheless, this study has the potential to enhance the current understanding of COVID-19 spreading and will help for the advancement of vaccination process of COVID-19. More over this study indicates that the COVID-19 containment policy i.e. lockdown adapted across the globe leads to reduced pollution level, improve climatic conditions in last few months. Such exceptional change has never been observed in nature before in short time duration. Therefore, such activity in future would be effective in sustainable development of nature.

## Data Availability

The digital dataset for the COVID-19 in India has been obtained from Ministry of Health and Family Welfare (MoHFW) [https://www.mohfw.gov.in/] and official website of covid-19 [https://www.covid19india.org/].

---

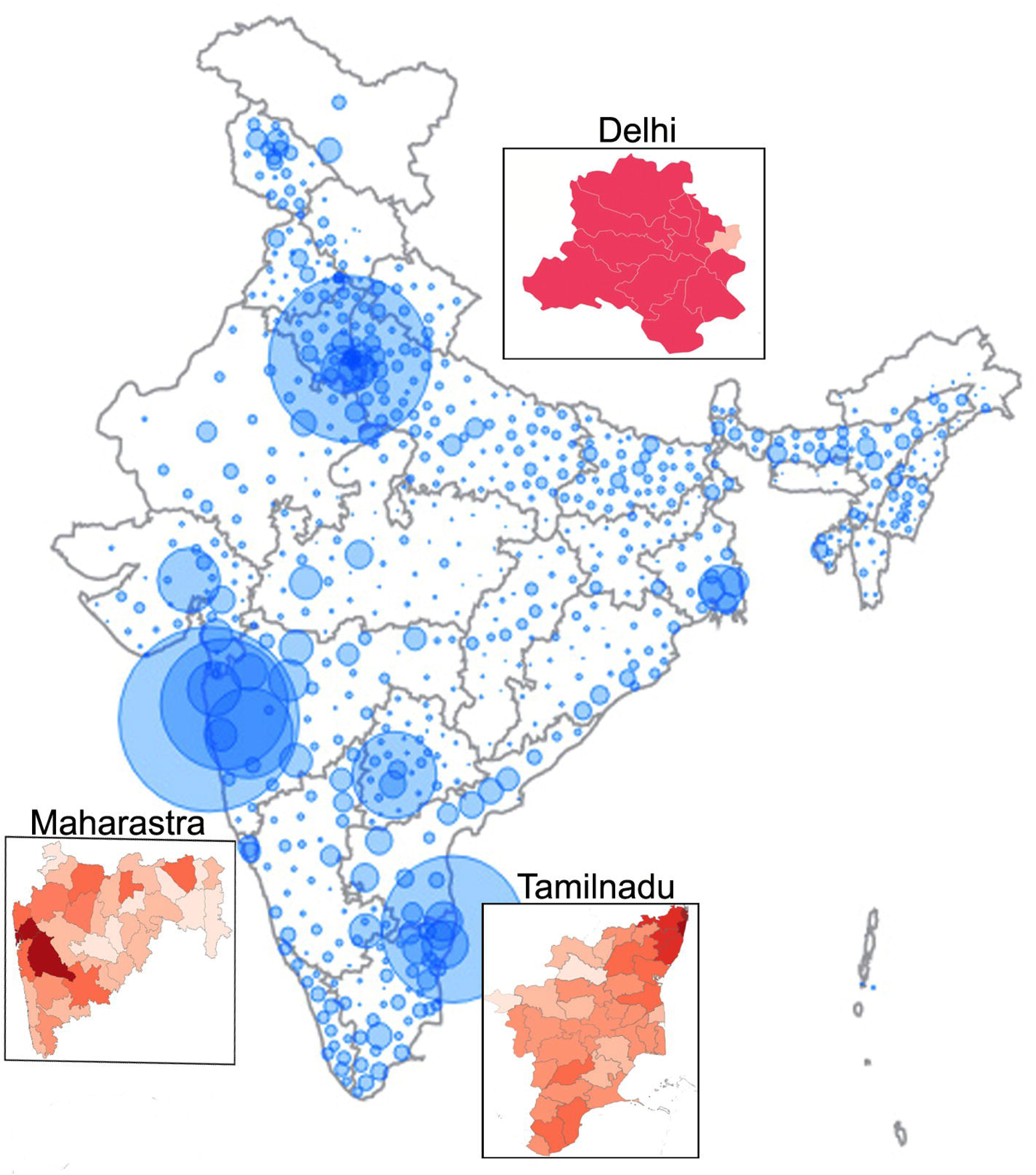

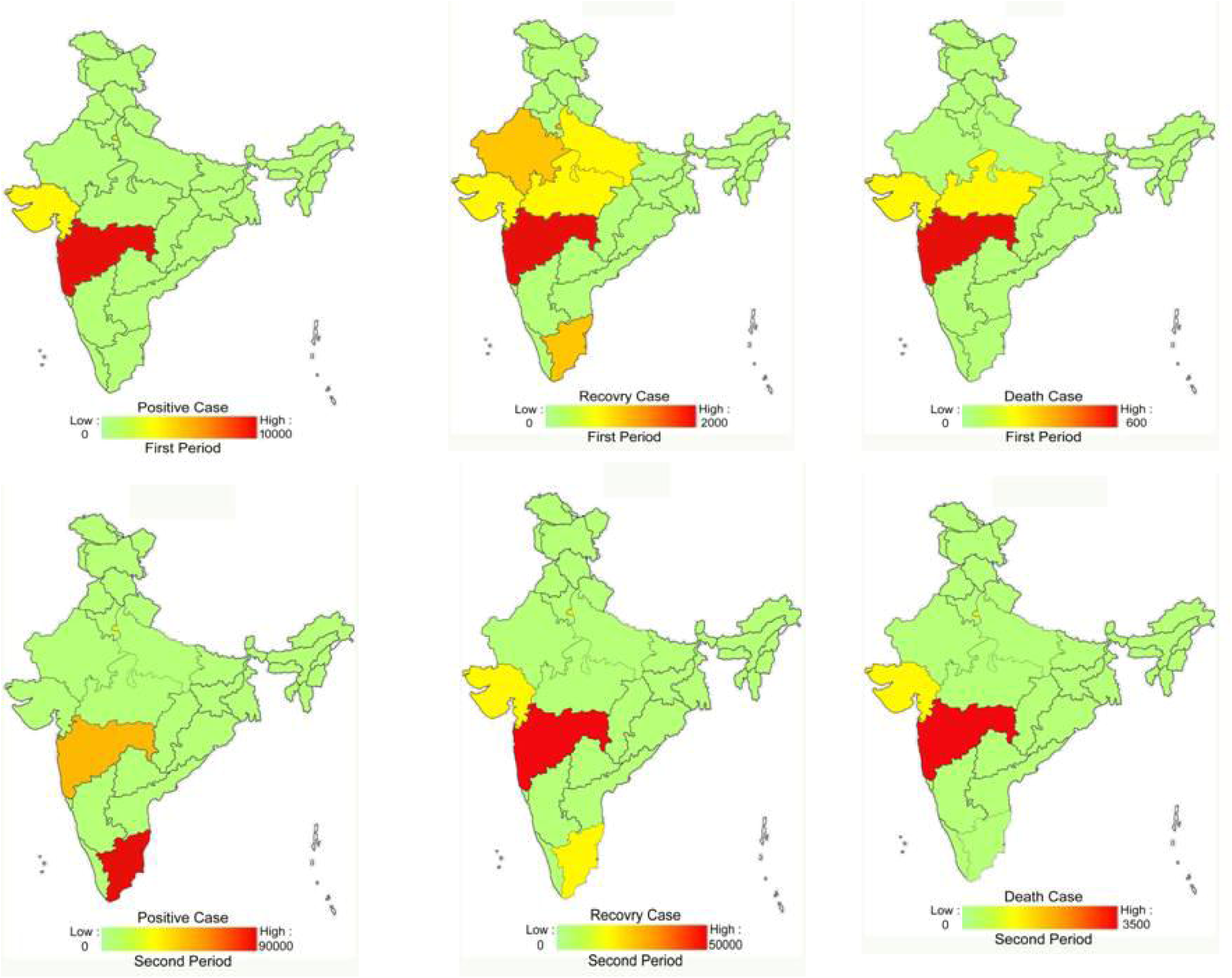

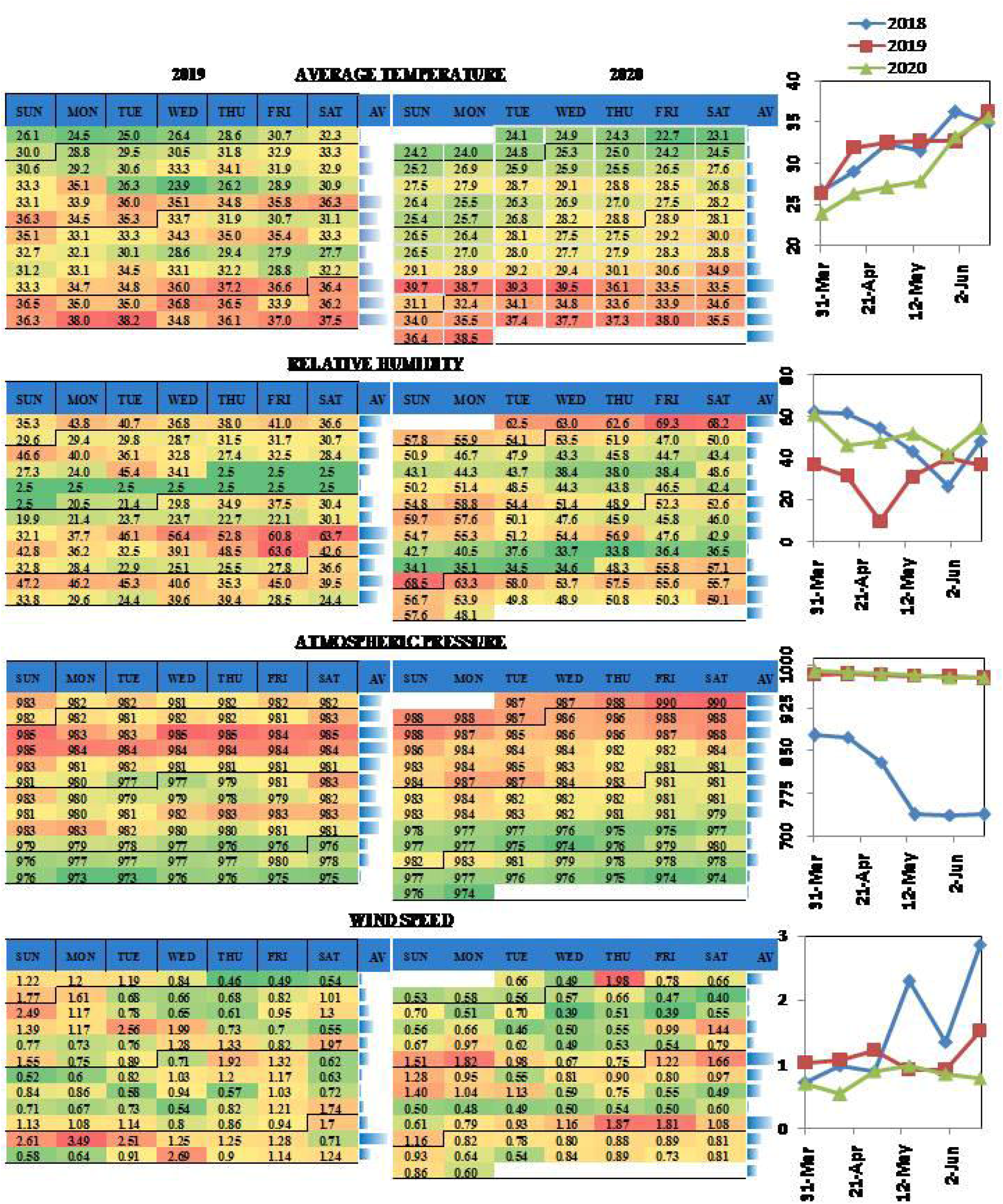

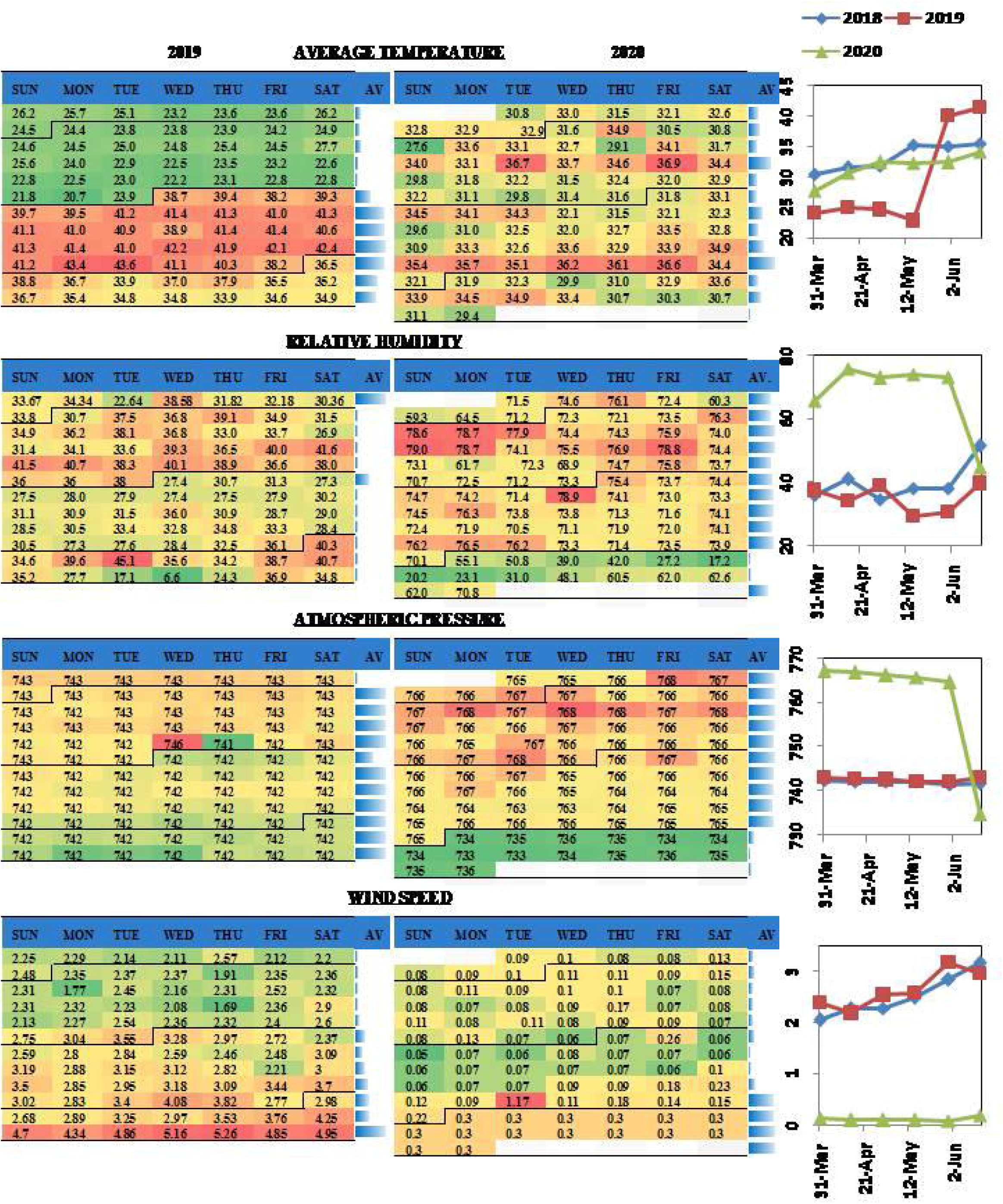

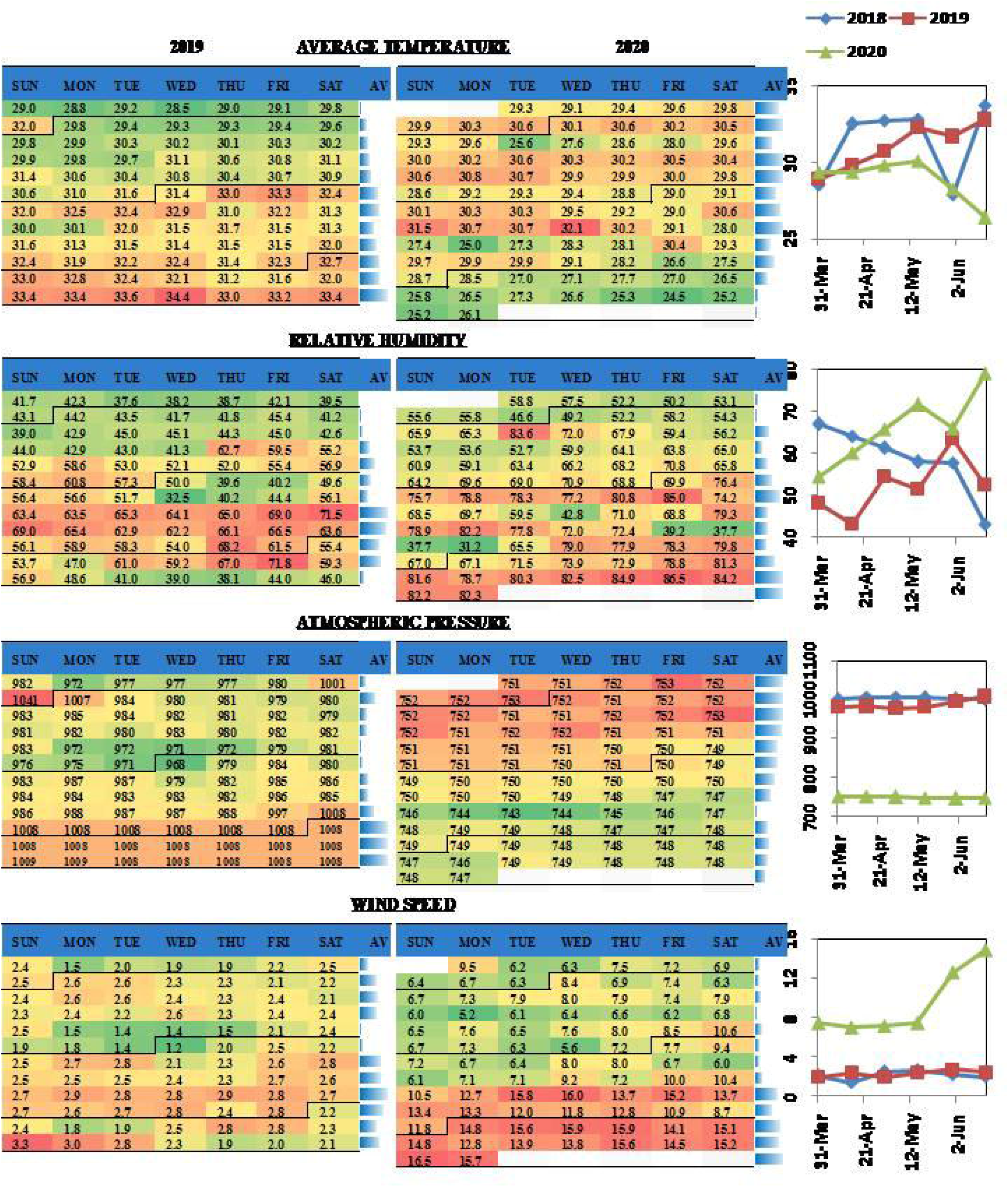

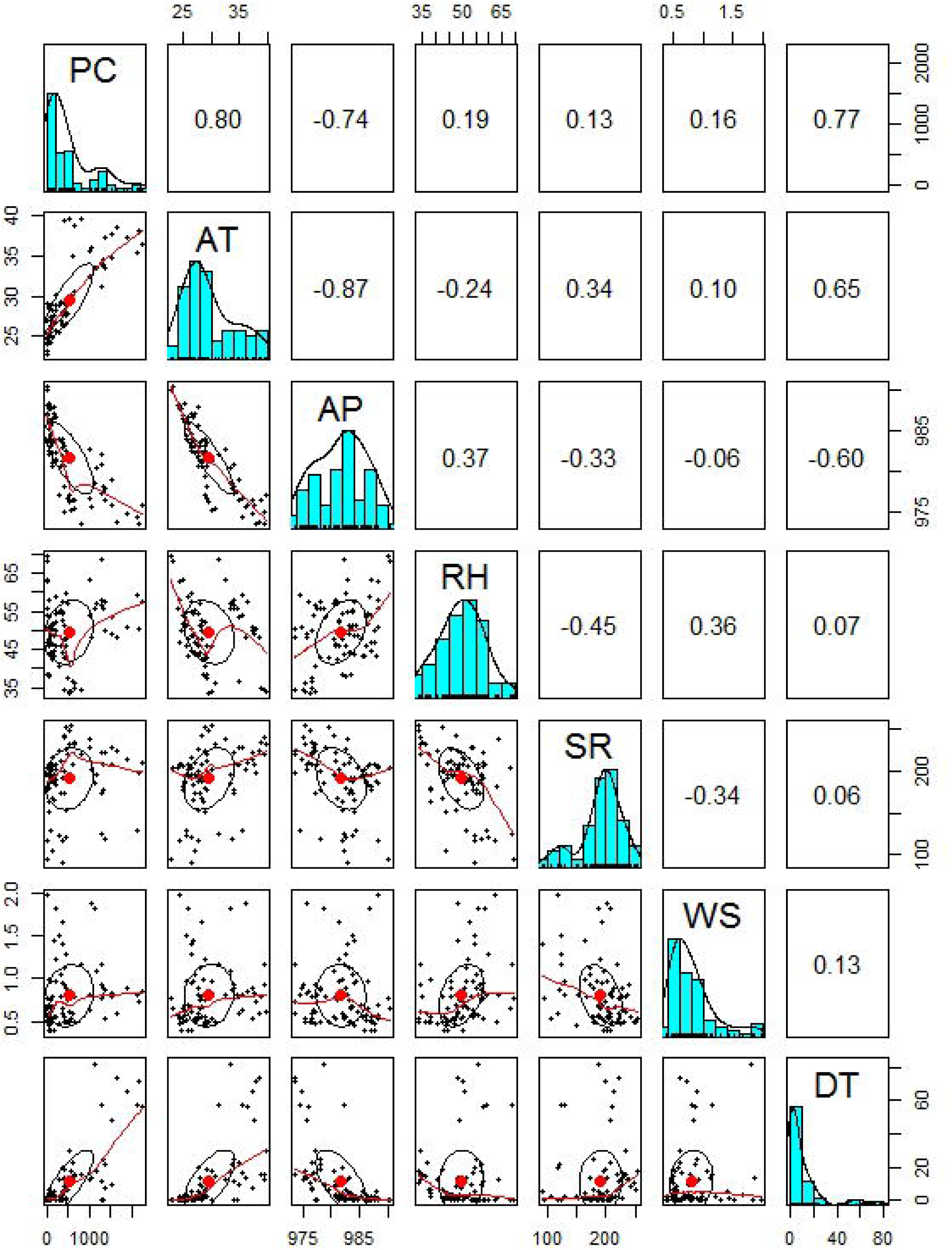

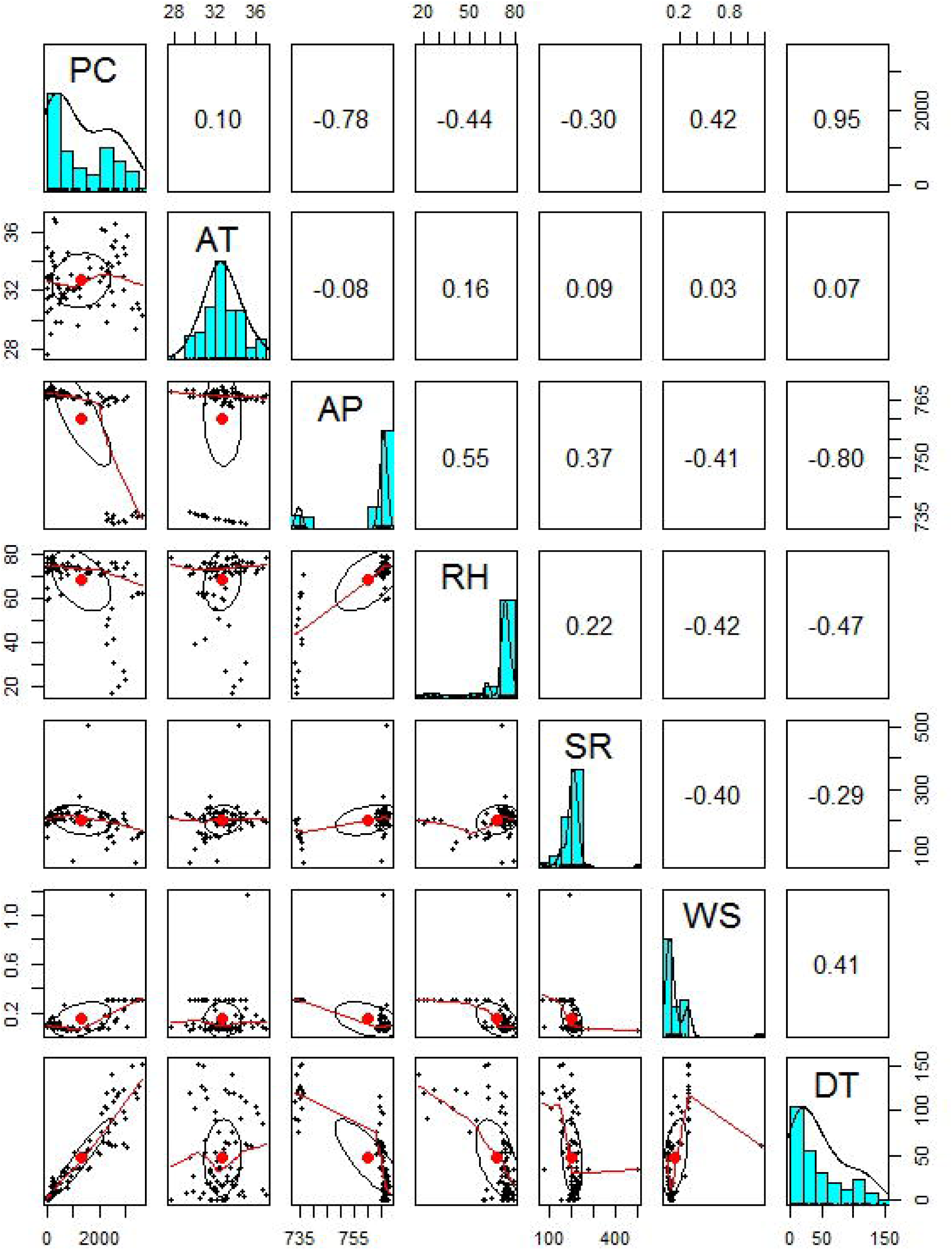

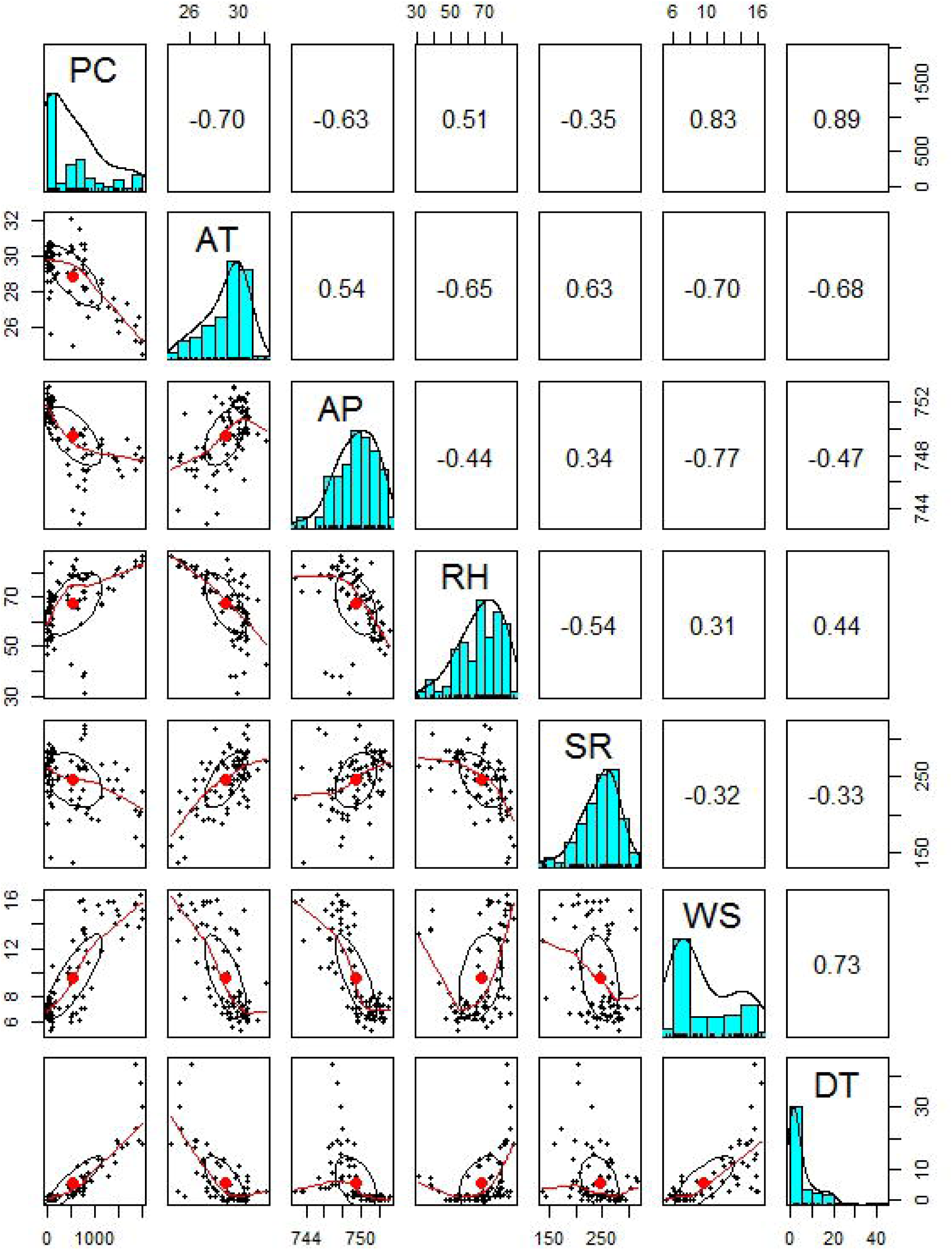

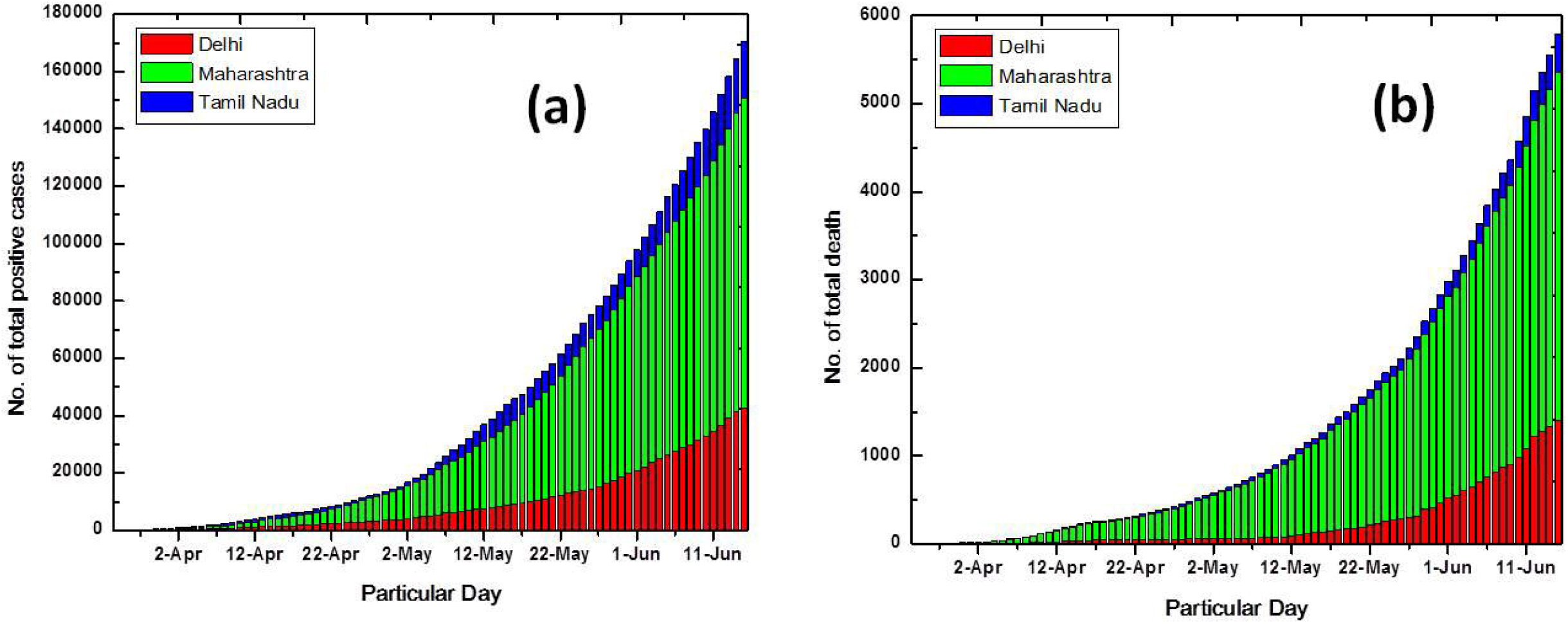

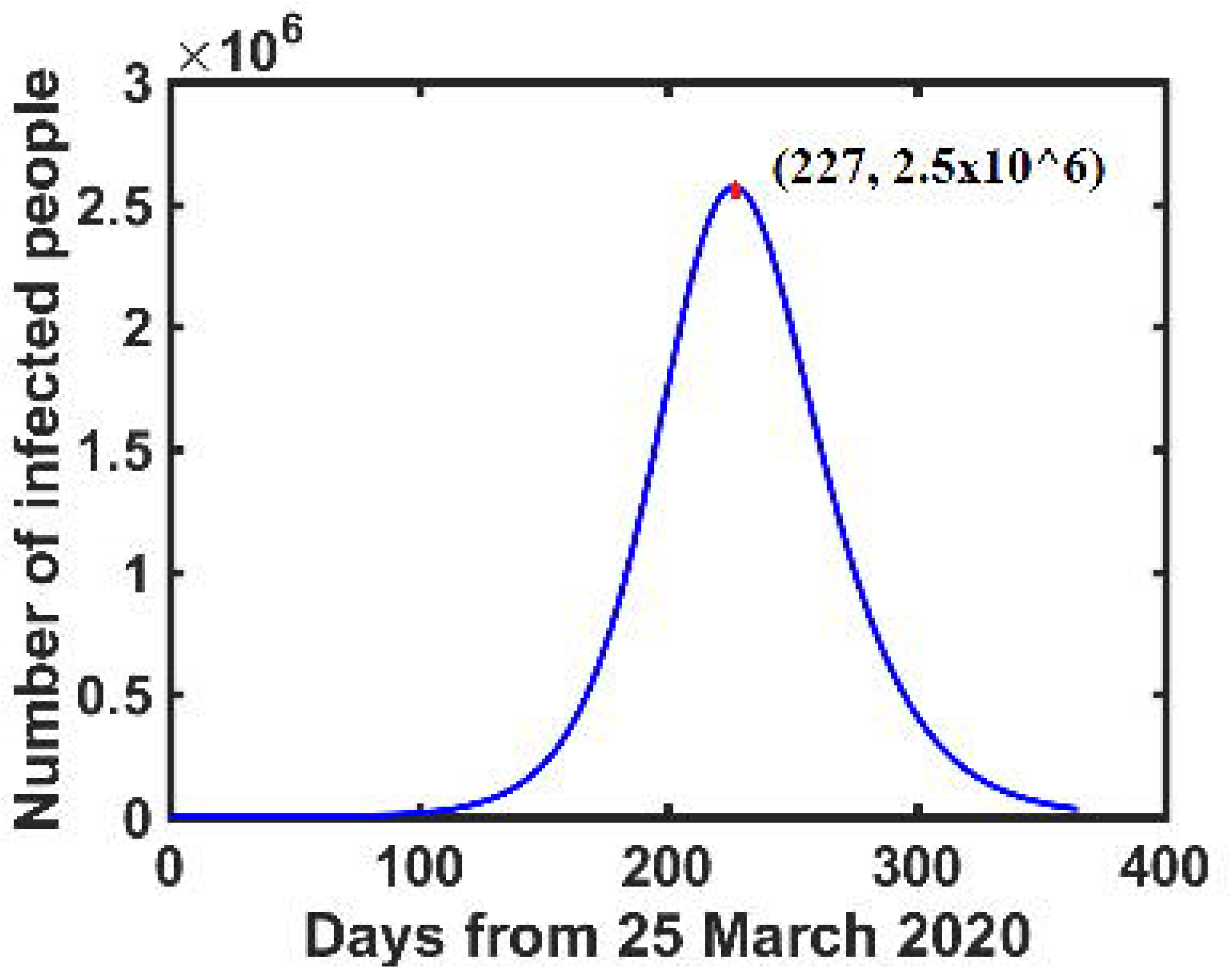

## References

Ahmadi, M., Sharifi, A., Dorosti, S., Jafarzadeh Ghoushchi, S., Ghanbari, N. 2020. Investigation of effective climatology parameters on COVID-19 outbreak in Iran. Science of The Total Environment 729:138705. doi.org/10.1016/j.scitotenv.2020.138705.

Briz- Redón, Á., Serrano-Aroca, Á. 2020. A spatio-temporal analysis for exploring the effect of temperature on COVID-19 early evolution in Spain. Science of The Total Environment 728: 138811. doi.org/10.1016/j.scitotenv.2020.138811.

Chen B, Liang H, Yuan X, Hu Y, Xu M, Zhao Y, et al. 2020. Roles of climaticconditions in COVID-19 transmission on a worldwide scale. doi.org/10.1101/2020.03.16.20037168

Cucinotta, D., Vanelli, M. 2020. WHO declares COVID-19 a pandemic. Acta bio- medica: Atenei Parmensis 91:157.

Dalziel, B.D.K, issler, S., Gog, J.R., Viboud, C., Bjørnstad, O.N., Metcalf, C.J.E., et al. 2018. Urbanization and humidity shape the intensity of influenza epidemics in U.S. cities. Science 362:75–79. doi:10.1126/science.aat6030.

Deepak, A.D., Hasan, KS., Joshua, L.M.D., Victor, N.F.D., David, A.M., and Erin, A.B. 2020. COVID-19 for the cardiologist: A Current Review of the Virology, Clinical Epidemiology, Cardiac and Other Clinical Manifestations and Potential Therapeutic Strategies. JACC; Basic to Translational science.

Guan, W. N i Z., Hu, Yu, Liang, W., Ou, C., He, J., Liu, L., Shan, H., Lei, C., Hui, D.S., Du, B.,Li, L., Zeng, G., Yuen, K.-Y., Chen, R., Tang, C., Wang, T., Chen, P., Xiang, J., Li, S., Wang,Jin-lin, Liang, Z., Peng, Y., Wei, L., Liu, Y., Hu, Ya-hua, Peng, P.,Wang, Jian ming, Liu, J.,Chen, Z., Li, G., Zheng, Z., Qiu, S., Luo, J., Ye, C., Zhu, S., Zhong, N. 2020. Clinical characteristics of 2019 novel coronavirus infection in China. medRxiv 2020.02.06.20020974.

Ge, X.-Y., Li, J.-L., Yang, X.-L., Chmura, A.A., Zhu, G., Epstein, 285 J.H., Mazet, J.K., Hu, B., Zhang, W., Peng, C., Zhang, Y.-J., Luo, C.-M., Tan, B., Wang, N., Zhu, Y., Crameri, G., Zhang, S.-Y., Wang, L.-F., Daszak, P., Shi, Z.-L. 2013. Isolation and characterization of a bat SARS-like coronavirus that uses the ACE2 receptor. Nature 289(503):535–538. doi:10.1128/JVI.00831-20.

Gupta, S., Raghuwanshi, G.S., Chanda, A. 2020. Effect of weather on COVID-19 spread in the US: A prediction model for India in 2020. Science of The Total Environment 728:138860. DOI: 10.1016/j.scitotenv.2020.138860.

Hänsel, S., Medeiros, D.M., Matschullat, J., Petta, R.A., de Mendonça Silva, I., 2016. Assessing Homogeneity and Climate Variability of Temperature and Precipitation. Series in the Capitals of North-Eastern Brazil. Front. Earth Sci. 4.

Huang, C., Wang, Y., Li, X., Ren, L., Zhao, J., Hu, Y., Zhang, L., Fan, G., Xu, J., Gu, X., Cheng,Z., Yu, T., Xia, J., Wei, Y., Wu, W., Xie, X., Yin, W., Li, H., Liu, M., Xiao, Y., Gao, H., Guo, L., Xie, J., Wang, G., Jiang, R., Gao, Z., Jin, Q., Wang, J., Cao, B. 2020. Clinical features of patients infected with 2019 novel coronavirus in Wuhan, China. The Lancet 395: 497–506. doi.org/10.1016/S0140-6736(20)30183-5.

Holshue, M.L., DeBolt, C., Lindquist, S., Lofy, K.H., Wiesman, J., and Bruce, H. 2020. First case of 2019 novel coronavirus in the United States. New England Journal of Medicine 382:929– 936. doi:10.1056/NEJMoa2001191.

Jaiswal, R.K., Lohani, A.K., Tiwari, H.L. 2015. Statistical Analysis for Change Detection and Trend Assessment in Climatological Parameters. Environ. Process 2:729–749. doi 10.1007/s40710-015-0105-3.

Jamil, T., Alam, I.S., Gojobori, T., Duarte, C. 2020. No Evidence for Temperature- Dependence of the COVID-19 Epidemic. medRxiv 2020.03.29.20046706.

Li, Q., Guan, X.,Wu, P., Wang, X., Zhou, L., Tong, Y., et al. 2020. Early transmission dynamics in Wuhan, China, of novel coronavirus–infected pneumonia. New England. Journal of Mediciane 382:1199–1207. doi:10.1056/NEJMoa2001316

M. F. Bashir, B. Ma, D. Bilal et al., 2020.Correlation between climate indicator and COVID-19 pandemic in New York, USA. Science of the Total Environment 728:138835.doi:10.1016/j.scitotenv.2020.138835

Ma Y, Zhao Y, Liu J, He X,Wang B, Fu S, et al.2020. Effects of temperature variation and humidity on the death of COVID-19 in Wuhan. Science of the total environment 724:138226. doi.org/10.1016/j.scitotenv.2020.138226

Mollalo, A., Vahedi, B., Rivera, K.M., 2020. GIS-based spatial modeling of COVID-19 incidence rate in the continental United States. Science of The Total Environment 728:138884. doi.org/10.1016/j.scitotenv.2020.138884

Perlman, S. 2020. Another decade, another coronavirus. New England. Journal of Mediciane 382:760–762. doi:10.1056/NEJMe2001126

Poole, L., 2020. Seasonal Influences on the Spread of SARS-CoV-2 (COVID19), Causality, and Forecastabililty (3-15-2020). Causality, and Forecastabililty (3-15-2020). (March 15, 2020).

Qi, H., Xiao, S., Shi, R., Ward, M.P., Chen, Y., Tu, W., Su, Q., Wang, W., Wang, X., Zhang, Z., 2020. COVID-19 transmission in Mainland China is associated with temperature and humidity: A time-series analysis. Science of The Total Environment 728, 138778.doi:10.1016/j.scitotenv.2020.138778.

Ramadhan Tosepu, Joko Gunawanb, Devi Savitri Effendy, La Ode Ali Imran Ahmad, Hariati Lestari, Hartati Bahar, Pitrah Asfian. 2020. Correlation between weather and Covid-19 pandemic in Jakarta, Indonesia. Science of the Total Environment 725:138436. doi.org/10.1016/j.scitotenv.2020.138436.

Rocklov, j., Sjodin, H. 2020. High population density catalysis the spreed of COVID-19. J Travel Med 27. doi:10.1093/jtm/taaa038.

Shi, P., Dong, Y., Yan, H., Li, X., Zhao, C., and Liu, W. 2020. The impact of temperature and absolute humidity on the coronavirus disease 2019 (COVID-19) outbreak evidence from China. medRxiv, doi.org/10.1101/2020.03.22.20038919.

Shi, Y., Ren, X., Niu, J., Zhu, W., Li, S., Luo, B., Zhang, K., 2020. Impact of climaticfactors on the COVID-19 transmission: A multi-city study in China. Science of The Total Environment 726, 138513. doi.org/10.1016/j.scitotenv.2020.138513.

Sajadi,M.M., Habibzadeh, P., Vintzileos, A., Shokouhi, S., Miralles-Wilhelm, F.,Amoroso, A. 2020. Temperature and Latitude Analysis to Predict Potential Spread and Seasonality for COVID-19. doi.org/10.2139/ssrn.3550308.

Sohrabi, C., Alsafi, Z., O’Neill, N., Khan, M., Kerwan, A., Al-Jabir, A., et al. 2020. World Health Organization declares global emergency: a review of the 2019 novel coronavirus (COVID-19). International Journal of Surgery 76:71–76. doi.org/10.1016/j.ijsu.2020.02.034.

Tan, J., Mu, L., Huang, J., Yu, S., Chen, B., Yin, J. 2005. An initial investigation of the association between the SARS outbreak and weather: with the view of the environmental temperature and its variation. J Epidemiol Community Health 59:186–192. doi:10.1136/jech.2004.020180.

Vandini, S., Corvaglia, L., Alessandroni, R., Aquilano, G.,Marsico, C., Spinelli, M., et al., 2013. Respiratory syncytial virus infection in infants and correlation with climaticfactors and air pollutants. Italian Journal of Pediatrics 39:1. doi:10.1186/1824-7288-39-1.

Wang, Y., Wang, Y., Chen, Y., and Qin, Q. 2020. Unique epidemiological and clinical features of the emerging 2019 novel coronavirus pneumonia (COVID- 19) implicate special control measures. Journal of medical virology. 92: 568–576. doi.org/10.1002/jmv.25748

Weisstein, Eric W. “SIR Model.” From MathWorld--A Wolfram Web.

Yuan, J., Yun, H., Lan, W., Wang, W., Sullivan, S.G., Jia, S., Bittles, A.H., 2006. A climatologic investigation of the SARS-CoV outbreak in Beijing, China. Am J Infect Control 34, 234–236. doi:10.1016/j.ajic.2005.12.006

Zhu, N., Zhang, D., Wang, W., Li, X., Yang, B., and Song, J. 2020. A novel coronavirus from patients with pneumonia in China, 2019. New England Journal of Medicine 382:727–733 doi:10.1056/NEJMoa2001017.

Zu, Z.Y., Jiang, M.D., Xu, P.P., Chen, W., Ni, Q.Q., and Lu, G.M. 2020. Coronavirus disease 2019 (COVID-19): A perspective from China. Radiology 200490.doi.org/10.1148/radiol.2020200490.

